# The time course of injury-risk after return-to-play in professional football

**DOI:** 10.1101/2023.09.25.23295972

**Authors:** Guangze Zhang, Michel Brink, Karen aus der Fünten, Tobias Trolß, Peter Willeit, Tim Meyer, Koen Lemmink, Anne Hecksteden

**Author notes:** <u>Corresponding author:</u> Anne Hecksteden Universität Innsbruck, Department of Sport Science, Chair of Sports Medicine Fürstenweg 176, 6020 Innsbruck, Austria; ORCID: 0000-0003-3390-9619.

## Abstract

**Background:** Injury risk in professional football is increased in the weeks following return-to-play (RTP). However, the time course of injury risk after RTP (the hazard curve) as well as its influencing factors are largely unknown. This knowledge gap, which is arguably due to the volatility of instantaneous risk when calculated for short time intervals, impedes on informed RTP decision-making and post-RTP player management.

**Objectives:** To characterize the hazard curve for non-contact, time-loss injuries after RTP in male professional football and investigate the influence of the severity of the index injury and playing position.

**Methods:** Media-based injury records from the first German football league were collected over four seasons as previously published. Time-to-event analysis was employed for non-contact, time-loss injury after RTP. The Kaplan-Meier survival function was used to calculate the cumulative hazard function, from which the continuous hazard function was retrieved by derivation.

**Results:** 1623 observed and 1520 censored events from 646 players were analyzed. The overall shape of the hazard curve was compatible with an exponential decline of injury risk, from an approximately two-fold level shortly after RTP towards baseline, with a half-time of about four weeks. Interestingly, the peak of the hazard curve was slightly delayed for moderate and more clearly for severe index injuries.

**Conclusions:** The time course of injury risk after RTP (the hazard curve) can be characterized based on the Kaplan-Meier model. The shape of the hazard curve and its influencing factors are of practical as well as methodological relevance and warrant further investigation.

**Summary box:** **What is already known on this topic -** As football players return to play after an injury, the risk of incurring a subsequent injury is high. With (event-free) time, this elevated risk returns to baseline. However, the shape of the risk-trajectory over time as well as its influencing factors are unknown.

**What this study adds -** This study characterizes the time course of injury risk after RTP by providing a continuous hazard curve. Moreover, differences in risk trajectories across severities of index injury and playing positions were investigated. **How this study might affect research, practice or policy -** An evidence-informed estimate of the excess injury risk still remaining at a certain time-point after RTP is of obvious use for RTP decision making and post-RTP player management. Moreover, the continuous hazard curves enable informed specification of follow-up period in epidemiological studies and verification of the proportional hazard assumption in data analysis.

## Introduction

Injuries are common in professional football players and seemingly unavoidable in their career.^1^ As players return to play (RTP) from a previous injury, the risk of subsequent injury is high^2^ and the time point at which the risk returns to baseline is largely unknown. Given the economic and competitive implications of injury burden, decisions on the timing of return to play and post-RTP player management are of particular importance for stakeholders (e.g., coaches, players and managers).^3^ Although there are attempts to determine the time frame within which injury risk is elevated^4–7^, the time course of the excess injury risk after RTP has yet to be uncovered.

Regarding the interval between RTP and subsequent injury, 50-80% of subsequent injuries are reported to occur within the first four weeks^4,7^. However, those figures are based on injury frequency and do not consider the fact that the number of players still at risk (i.e., the cohort that have not sustained a subsequent injury) also drops as time goes by because each injured player reduces this group. A decreasing number of players at risk would lead to a decline of injury occurrence even with constant injury risk. Time-to-event analysis^8^ (survival analysis) has been widely employed to investigate the occurrence and timing of events in epidemiology^9^ and beyond^10^. When applied to injury occurrence, time-to-event analysis (e.g., using the Kaplan-Meier method) enables incorporating the decreasing number of players at risk. Previous studies have generally quantified injury risk after RTP in coarse time intervals of 2 to 8 weeks^4,7,11^. In such a discrete time framework^12^, injury risk for a specific time interval can be conveniently estimated by dividing the number of injuries by the number of players still at risk^12^. However, discrete hazard estimation with a coarse time metric (e.g., months) may be inadequate to capture the fast changes in injury risk after RTP. The key challenge for a fine-grained time scale (e.g., days) is the volatility of hazard estimates due to the low number of injuries per time interval (including intervals without injury at all). As a consequence, the hazard function cannot be directly estimated in a continuous time framework^12^. The present work explores the application of an established statistical solution^12^ within the field of injury risk after RTP.

In the analysis of injury occurrences and RTPs over a football career, two classes of time-intervals must be distinguished. The interval between an (index) injury and RTP (i.e., RTP time) and the interval between RTP and the subsequent injury. While the present work will be focusing on the latter, RTP time is also of interest for the following two reasons. Firstly, according to the time-loss concept, time to RTP is used as an indicator of injury severity^13^ with implications for post-RTP injury risk^14^. The respective consensus has been widely applied^7,15^. Secondly, RTP time relative to the initial diagnosis of the injury could allow an estimation on rehabilitation (in)adequacy which also influences injury risk after RTP^6,16^. In addition, the playing position is also an established influencing factor of injury risk within and beyond the context of RTP.^1^

In existing time-to-injury analysis in football, Cox regression models have been widely used to estimate discrete hazard ratios for supposed risk factors^16^, including in the RTP context^7^. However, a fundamental assumption of the Cox model (proportional hazard) has not been verified so far. That is, that hazard functions for these risk factors are proportional (parallel hazard curves over time^17^) and their association can therefore be summarized as one common hazard ratio.

Contact injuries are relatively unpredictable in football.^18^ Therefore, the present work aims to characterize the continuous-time hazard curve of non-contact injuries after RTP in professional football. Moreover, variations of the continuous hazard curve among severities of index injury and playing positions will be investigated. The hazard curves may also verify whether the proportional assumptions are met.

## Methods

### Dataset

The analyzed data included four seasons (2014/15 to 2017/18) of media-based injury records in the 1st male German football league, a subset of the dataset used by aus der Fünten et al^19^. Neither research ethics board approval nor a trial registration was required as all data were collected from publicly available sources. The primary data source was the online version of the sport-specific journal “kicker Sportmagazin^TM^”^20–22^ complemented by further publicly available resources. Injury data collection followed the Fuller consensus statement on football injury studies^13^. All injury records were subsequently examined by an independent orthopedic doctor for medical plausibility.^19^

### Equity, diversity and inclusion statement

The focus of this work is on male professional football. While the specific results are presumably dependent on discipline, performance level and sex, the method presented may be applied in other settings and populations.

### Data processing

The severity of index injuries was categorized according to the time loss concept: minimal (1-3 days), mild (4-7 days), moderate (8-28 days), and severe (>28 days).^13^ The playing position was considered as players’ main position when the subsequent injury occurred, including goalkeeper, defender, midfielder, and forward.

As contact and non-contact injuries can equally impact on players’ physical condition and subsequently influence the injury risk after RTP, both categories are considered for the index injury. However, given the unpredictability of physical contact in football, only non-contact, time-loss injuries were considered as subsequent injuries. For each player, the first injury in record was considered as the index injury of the following one, the second injury as the index injury of the third one, and so on. RTP in this study was defined as a full return to training and competition^7^.

### Censoring

Censoring refers to abbreviated length of follow-up due to the end of the follow-up period or reasons other than the target event. Four football seasons were segmented by the date of the last official match for corresponding season, 23^rd^ May for 2014/15 season, 14^th^ May for 2015/16 season, 20^th^ May for 2016/17 season, and 12^nd^ May for 2017/18 season. Given that training and match exposure as well as the recording of minor injuries during the summer break might differ from in-season, cases that did not incur a subsequent injury in the same season as RTP were censored at the end of season (date of last match, cp. above and Fig. 1). As this study mainly focused on the occurrence of non-contact injury after RTP, contact-related subsequent injuries equally led to censoring (Fig. 1). Altogether, a subsequent injury was confirmed as an observed event only when it was observed in both categories (i.e., non-contact subsequent injury occurring in the same season as RTP).

**Fig. 1:**
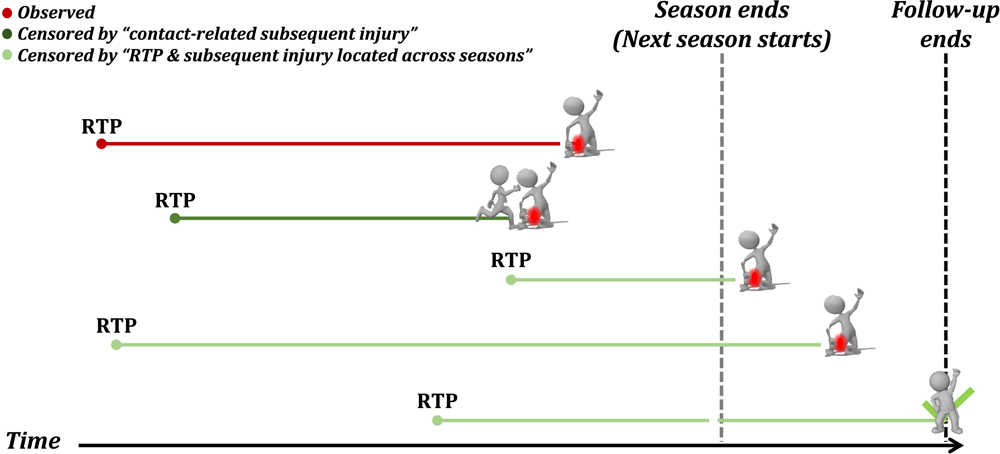
Two strategies of censoring observations. The example at the bottom is not subsequently injured until the end of follow-up.

### Hazard function

Fig. 2 illustrates the steps to derive the continuous hazard function. First, the dataset with censoring information was used to fit a Kaplan-Meier (KM) model^8^. Due to the fine-grained time metric (day), the number of observed events might be trivial for individual time units, which makes the discrete-time KM hazards too volatile to be meaningful. By contrast, at each time c_j_, the cumulative hazard function ^w^Cc_j_) can be derived through an established mathematical relationship (see equation 1) from the KM survival function s^-^_KM_ Cc).^12^ The instantaneous risk is the change in cumulative hazard from one time unit (day) to the next, that is, the local slope of the cumulative hazard function. The cumulative hazard function provides a pivot to retrieve continuous hazards as its first derivative.^12^ To simplify calculation, cumulative hazards and time, as response and explanatory variables respectively, were used to fit a polynomial (10th degree) regression model. Subsequently, predictions were made for successive days. The rate of change in predicted cumulative hazards was then calculated as the continuous hazard function (i.e., the risk of subsequent injury).^12^ Note that only cumulative hazards from the first 100 days after RTP were used to fit the polynomial.

**Fig. 2:**
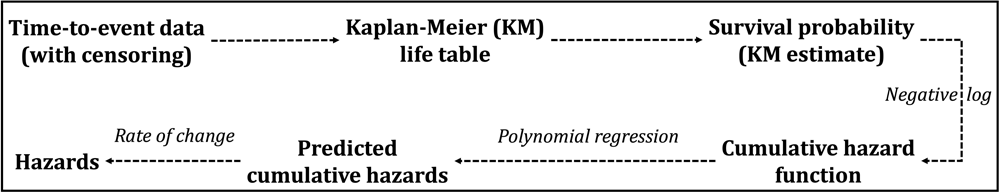
Retrieving the hazard function on continuous time.

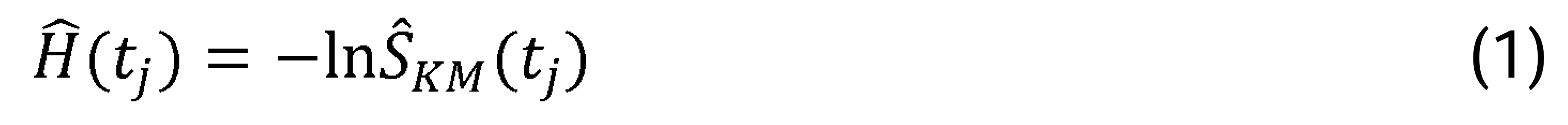

A linear interpolation approach ^8^ was applied to estimate the median survival time r, as shown in equation 2 where m represents the time interval when the sample survival function is just above 0.5.

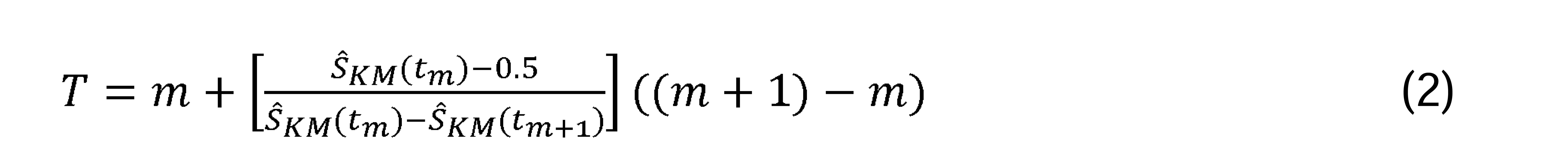

Note that the analysed dataset features a hierarchical data structure. As players with more frequent injuries (and therefore RTPs) contribute more data points, these individuals have a disproportionately high impact on the hazard function, leading to potential bias. Aiming to expose the general analytical pipeline for deriving the continuous hazard function as transparently as possible, the nesting of RTP episodes within players is not considered in the above analysis. However, in supplement, we illustrate and discuss two potential options for mitigating this issue while still avoiding advanced modelling techniques.

## Results

### Epidemiology of subsequent injuries

Within the four seasons, 822 players incurred 4065 injuries, with a total of 3143 subsequent injuries from 646 players. 674 (21.4%) subsequent injuries occurred across the end of season, which in conjunction with contact-related injuries (*n*=1102, 35.1%) resulted in 1520 censored cases and 1623 observed subsequent injuries.

77% (*n*=2406) of all subsequent injuries and 83% (*n*=1343) of observed subsequent injuries were sustained during the first 100 days after RTP. The median survival time was 84 days after RTP (Fig. 3). Fig. 4 showed that the number of players still at risk steadily fell over the post-RTP period. Observed subsequent injuries presented a similar overall pattern, however, with fluctuation.

**Fig. 3:**
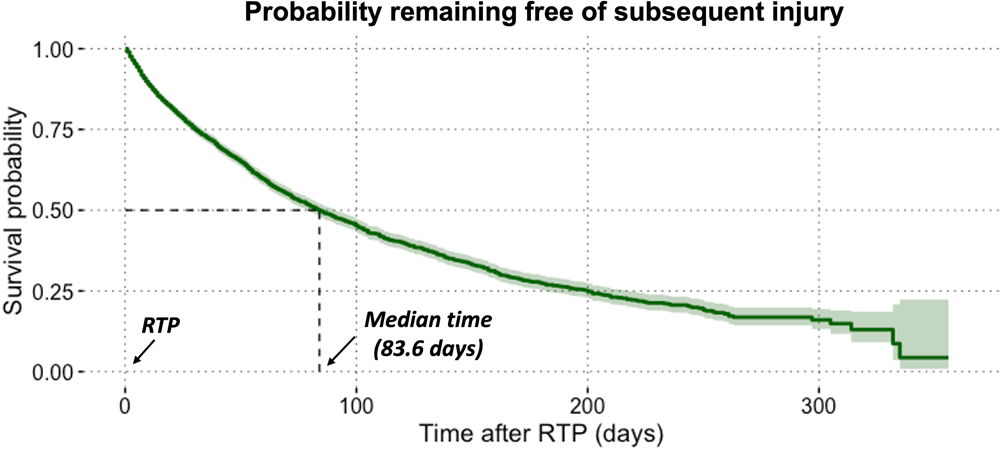
Kaplan-Meier estimates of continuous-time survivor function with 95% confidence interval and median survival time.

**Fig. 4:**
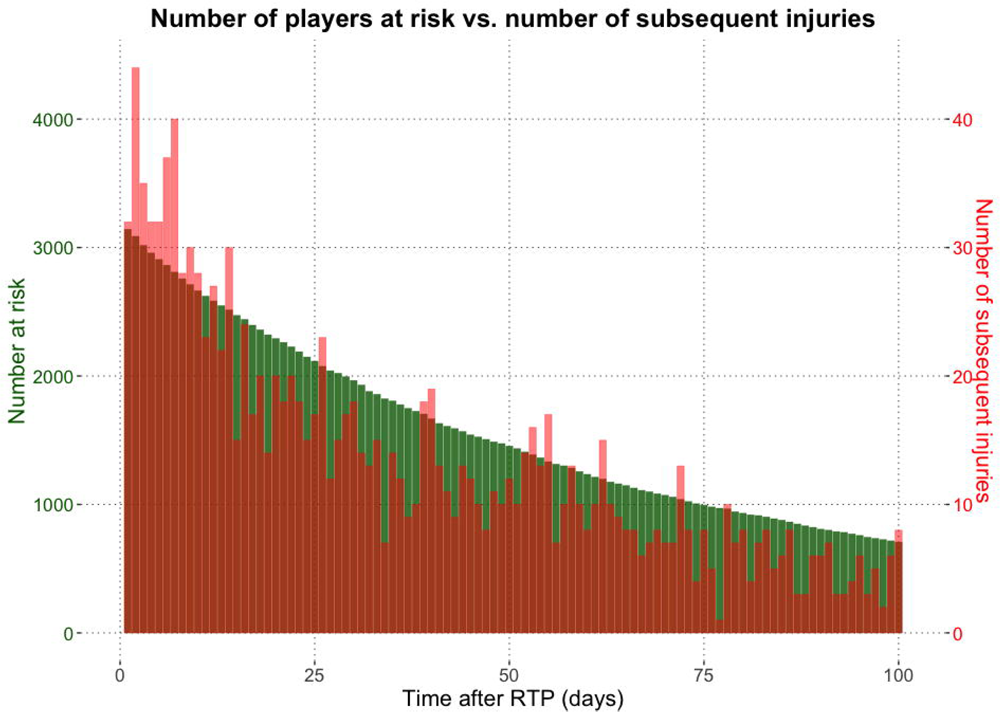
The count of players still at risk (green bar) and observed subsequent injuries (red bar) at time course after RTP.

Among all observed subsequent injuries, minimal, mild, moderate, and severe index injuries contributed 38.9%, 20.2%, 24.5%, and 16.3%, respectively. The respective proportions of severity categories were similar across playing positions (Fig. 5). With respect to playing positions, 551 (including 278 observed) subsequent injuries were suffered by 141 forwards, 1406 (696 observed) subsequent injuries by 272 midfielders, 1051 (582 observed) subsequent injuries by 236 defenders, 135 (67 observed) subsequent injuries by 41 goalkeepers.

**Fig. 5:**
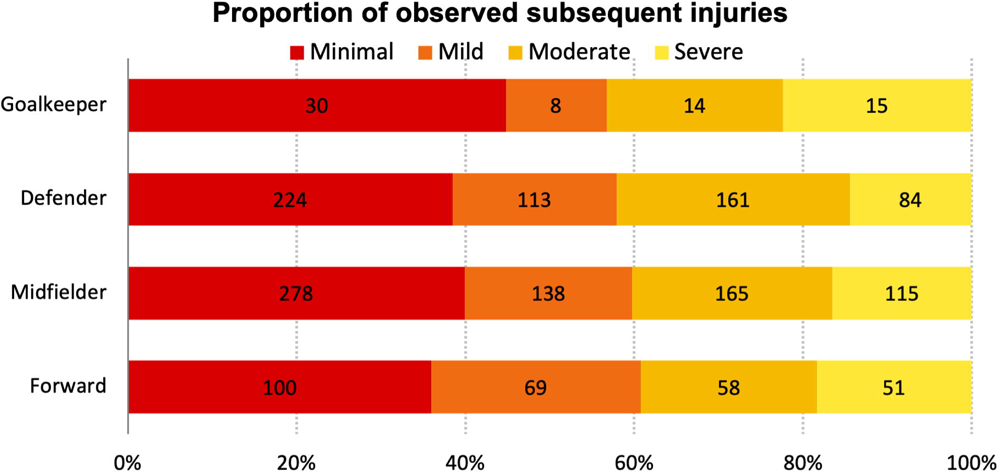
Distribution of observed subsequent injuries after different severities of index injury across playing positions.

### Risk of subsequent injury (hazard curve)

Overall, as players returned to play, the risk of non-contact subsequent injury was about two times higher than the baseline. Across all analyzed events, the shape of the hazard curve is compatible with an exponential decay of excess risk, which diminished by half after approximately 25 days and levels off afterwards (Fig. 6a). There is a larger relative change over time when analyzing injury frequencies (the red bars in Fig. 4) vs. hazards (Fig. 6a) which take the number of players at risk into account.

**Fig. 6:**
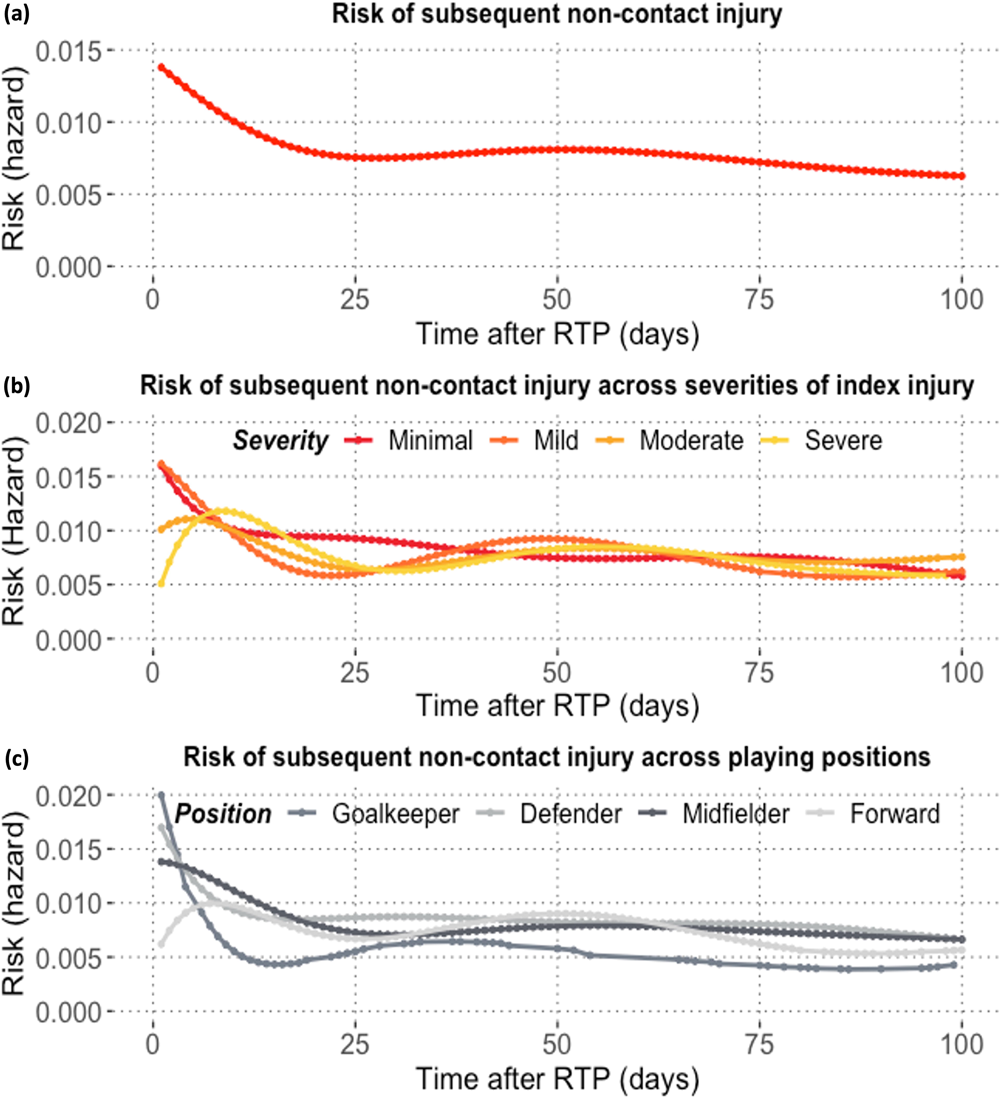
The time course of a) non-contact injury risk after RTP; non-contact subsequent injury risk across b) severities of index injury, and c) playing positions.

As shown in Fig. 6b, the shape of the hazard curve differs across severities of the index injury. For minimal and mild index injuries, the “exponential” pattern holds. A minor risk increment over the first five days was observed for moderate index injuries and RTP from severe injuries was followed by a significantly increasing injury risk within the first 10 days, which thereafter remained relatively high.

Goalkeepers, midfielders, and defenders faced a decreasing risk of non-contact injuries in the first four weeks as returning to full football activity (Fig. 6c). The greatest plunge of injury risk was seen in goalkeepers, with an approximate decline by 75% in the two weeks after RTP. In contrast, forwards experienced a slightly increasing risk of getting injured again after RTP which fluctuated over the post-RTP period.

## Discussion

The primary aim of this study was to derive and investigate the hazard curves for subsequent non-contact injuries after RTP in professional football. The overall shape of the hazard curve followed the expected pattern of an exponential “decay” of excess risk towards baseline. Importantly, in contrast to previous studies^4,7^ looking into the timing of subsequent injuries after RTP, the current study determined a continuous hazard function, thereby avoiding overestimation of changes in injury risk. With respect to the risk across severities of index injury and playing positions, as opposed to other groups in each category, a delayed maximum of hazard was found for those returning from moderate and severe index injuries and for forwards. This is indicative of a period and a group of players which need particular attention. Hazards appeared not to be proportional for these groups.

Bengtsson et al.^2^ reported an increased injury rate in the first match appearance after RTP compared to the average seasonal match injury rate for all injuries (46.9 vs 25.0/1000 hours) and for muscle injuries (24.6 vs 9.5/1000LJhours) when injury risk was simply calculated by dividing the number of injured players by the total number of players. In a 3-year follow-up study^23^, 6 (6.7%) players after ACL reconstruction suffered complications (five re-ruptures and four other knee injuries) between return-to-training and the first match. While, similarly, Orchard et al.^5^ found that players in Australian Football League faced the highest injury risk during the first week after RTP, they also reported that risk was still increased during the following weeks. This could be explained by still ongoing muscle regeneration after a rehabilitation of weeks^24^. Nevertheless, these findings were obtained from incidence rates averaged over a certain period and critically depended on the follow-up duration, thereby ignoring the potentially time-varying distribution of injury over time^17,25^. The current time-to-event analysis presented the continuous hazard of non-contact injuries after RTP and takes into account its time-varying characteristics.

While in the current study, the risks of subsequent injury steadily declined after returning from minimal and mild index injuries, rehabilitation adequacy for these cases should not be overlooked in practice^26^. Ekstrand and Gillquist^11^ reported that a minimal or mild injury could be followed by a more severe subsequent injury. Severe injuries are usually less common in football but cause longer absence times.^15^ The delayed peak of injury risk after returning from severe index injuries may be due to larger tissue damage, for example, damage of nerves and impaired proprioception. This might require longer time for regeneration. De-training effects after long absence and immobilization due to severe injuries could also undermine players’ muscle mass^27^, proprioception^15^, and cardiovascular capacity^28^. Players returning from severe injuries may play with greater care in the first days. After several regular training sessions, their confidence may be restored sooner than actual athletic capacity, which in conjunction with the expectation of proving themselves may lead to the delayed peak of injury risk. Thus, players returning from severe injuries warrant further attention and monitoring over the following weeks.

The trajectory of non-contact injury risk after RTP differed across playing positions. This could correspond with the fact that different roles were exposed to various intensities of physical contact in training and match^29,30^. Forwards faced a delayed peak shortly after RTP with some fluctuations over the post-RTP period. Carling et al^1^ similarly found that center-forwards sustained a higher incidence of recurrent muscle strains than other positions. Comparable results were reported for groin injuries.^31^ Nevertheless, findings are inconsistent across studies for an association between playing positions and injury risk.^32^ Some circulation of playing positions in modern football may explain the inconsistency. Classification of playing positions in this type of study may need further discussion.

In this study, the characterization of hazard curves enables a direct, visual assessment of the proportional hazard assumption. Importantly, hazard curves differed in overall shape and scale across severities of index injury and playing positions. This result points to the necessity of explicitly verifying the proportional hazard assumption for any given setting. The direct visual assessment of the quantity of interest could be a pragmatic approach especially when working with limited sample sizes^33^. Previously, more indirect visualizations have been used to examine this assumption, such as survival function against time, cumulative hazard versus time, log (cumulative hazard) versus log (time), or Schoenfeld residuals versus log (-survival function) ^9,34^. For example, Della Villa et al^35^ examined the proportional hazards assumption with Schoenfeld residuals when investigating potential factors associated with second ACL injuries, where the assumption was globally met for all candidate risk factors. Again, sample size and statistical power should be considered for the specific case.

### Strengths, limitations, and future directions

For the first time this study used time-to-event analysis to characterize the continuous hazard curves for subsequent non-contact injuries in football. However, it should be noted that the retrospective, media-based data set is associated with some limitations. In prospective follow-up studies, exposure hours (or exposure load) after RTP could provide more accurate insights into the time course of injury risk after RTP compared to days after RTP as it has been utilized here. Moreover, future research could include other influencing factors such as injury history (e.g., frequency of previous injuries)^36^, rehabilitation adequacy^14^, location (e.g., the body region^36,37^ and affected tissue^6,15^) and type^1,16^ of index injury. Of note, the findings from male professional football may not be applicable to other playing levels or female football, nor to other sports.

Finally, as aforementioned in the methods section, the main analysis did not consider the nesting of events within individuals. It has to be kept in mind that this might lead to bias because frequent injury occurrence leads to overrepresentation of episodes from the concerned player in the dataset and is, at the same time, plausibly associated with shorter time intervals (injury severity and time between RTP and subsequent injuries). Respecting the proof-of-concept character of this work, we consciously opted to focus on exposing the main analytical strategy. However, in the supplementary document we illustrate two potential solutions which still avoid advanced modeling technique (a) randomly up-sampling on the individual level within each season to the maximum number of RTPs per player in the corresponding season; (b) including only the first RTP of each player within each season (as a form of down-sampling). Both methods operate on the level of data processing without requiring alterations of the main analytical proceedings presented in the methods section. Importantly, all three analytical options result in a similar overall shape of the hazard curve.

## Conclusions

Through time-to-event analysis, this study determined the continuous hazard curve of non-contact injuries after RTP, which was two times higher at the day of RTP than the baseline level. One month follow-up after RTP is reasonable to capture most “surplus” risk of subsequent non-contact injury while avoiding excessive effort as well as “dilution” with injuries unrelated to RTP. The severity of index injury and playing position impact on the time course of the non-contact injury after RTP, resulting in a severity-dependent delay of the peak hazard.

## Practical implication

Post-RTP player management benefits from a valid estimate of the remaining excess injury risk as time elapses. This study demonstrates how to derive a continuous time hazard curve to support such decision making. However, replication and further investigation are warranted before applying our specific results in practice.

## Declarations

### Author contributions

All authors contributed to the drafting, writing, and editing of this article. All authors have read and approved the final manuscript.

## Supporting information

"supplement" at line 168, and "supplementary document" at line 301

## Data Availability

All data produced in the present study may be available upon reasonable request to the authors

## References

1. Carling C, Le Gall F, Orhant E. A four-season prospective study of muscle strain reoccurrences in a professional football club. Res Sports Med. 2011;19(2):92–102.

2. Bengtsson H, Ekstrand J, Waldén M, Hägglund M. Few training sessions between return to play and first match appearance are associated with an increased propensity for injury: a prospective cohort study of male professional football players during 16 consecutive seasons. British journal of sports medicine. 2020;54(7):427–432.

3. Yung KK, Ardern CL, Serpiello FR, Robertson S. A Framework for Clinicians to Improve the Decision-Making Process in Return to Sport. Sports Med Open. 2022;8(1):52.

4. Wangensteen A, Tol JL, Witvrouw E, et al. Hamstring Reinjuries Occur at the Same Location and Early After Return to Sport: A Descriptive Study of MRI-Confirmed Reinjuries. Am J Sports Med. 2016;44(8):2112–2121.

5. Orchard J, Best TM. The management of muscle strain injuries: an early return versus the risk of recurrence. Clinical Journal of Sport Medicine. 2002;12(1):3–5.

6. Gajhede-Knudsen M, Ekstrand J, Magnusson H, Maffulli N. Recurrence of Achilles tendon injuries in elite male football players is more common after early return to play: an 11-year follow-up of the UEFA Champions League injury study. Br J Sports Med. 2013;47(12):763–768.

7. Hagglund M, Walden M, Ekstrand J. Lower reinjury rate with a coach-controlled rehabilitation program in amateur male soccer: a randomized controlled trial. Am J Sports Med. 2007;35(9):1433–1442.

8. Miller R. Survival analysis. New York: Wiley; 1981.

9. Kuitunen I, Ponkilainen VT, Uimonen MM, Eskelinen A, Reito A. Testing the proportional hazards assumption in cox regression and dealing with possible non-proportionality in total joint arthroplasty research: methodological perspectives and review. BMC Musculoskelet Disord. 2021;22(1):489.

10. Morrison J. Introduction to survival analysis in business. Journal of Business Forecasting Methods and Systems. 2004;23(1):18–22.

11. Ekstrand J, Gillquist J. Soccer injuries and their mechanisms: a prospective study. Medicine and science in sports and exercise. 1983;15(3):267–270.

12. Singer JD, Willett JB. Applied longitudinal data analysis: Modeling change and event occurrence. Oxford university press; 2003.

13. Fuller CW, Ekstrand J, Junge A, et al. Consensus statement on injury definitions and data collection procedures in studies of football (soccer) injuries. Br J Sports Med. 2006;40(3):193–201.

14. Murphy DF, Connolly DA, Beynnon BD. Risk factors for lower extremity injury: a review of the literature. Br J Sports Med. 2003;37(1):13–29.

15. Ekstrand J, Krutsch W, Spreco A, et al. Time before return to play for the most common injuries in professional football: a 16-year follow-up of the UEFA Elite Club Injury Study. Br J Sports Med. 2020;54(7):421–426.

16. Hägglund M, Waldén M, Ekstrand J. Previous injury as a risk factor for injury in elite football: a prospective study over two consecutive seasons. British Journal of Sports Medicine. 2006;40(9):767.

17. Hernan MA. The hazards of hazard ratios. Epidemiology. 2010;21(1):13–15.

18. Hecksteden A, Schmartz GP, Egyptien Y, Aus der Funten K, Keller A, Meyer T. Forecasting football injuries by combining screening, monitoring and machine learning. Sci Med Footb. 2022:1–15.

19. Aus der Funten K, Tross T, Hadji A, Beaudouin F, Steendahl IB, Meyer T. Epidemiology of Football Injuries of the German Bundesliga: A Media-Based, Prospective Analysis over 7 Consecutive Seasons. Sports Med Open. 2023;9(1):20.

20. Werner K, Clemens M, Volker K, et al. High return to competition rate following ACL injury – A 10-year media-based epidemiological injury study in men’s professional football. European Journal of Sport Science. 2019;20:1–15.

21. Krutsch W, Memmel C, Alt V, et al. Timing return-to-competition: a prospective registration of 45 different types of severe injuries in Germany’s highest football league. Arch Orthop Trauma Surg. 2022;142(3):455–463.

22. Florian B, Karen aus der F, Tobias T, Claus R, Tim M. Head injuries in professional male football (soccer) over 13 years: 29% lower incidence rates after a rule change (red card). British Journal of Sports Medicine. 2019;53(15):948.

23. Waldén M, Hägglund M, Magnusson H, Ekstrand J. ACL injuries in men’s professional football: a 15-year prospective study on time trends and return-to-play rates reveals only 65% of players still play at the top level 3 years after ACL rupture. British journal of sports medicine. 2016;50(12):744–750.

24. Huard J, Li Y, Fu FH. Muscle injuries and repair: current trends in research. JBJS. 2002;84(5):822–832.

25. Dunn OJ, Clark VA. Basic statistics: a primer for the biomedical sciences. John Wiley & Sons; 2009.

26. Woods C, Hawkins R, Hulse M, Hodson A. The Football Association Medical Research Programme: an audit of injuries in professional football—analysis of preseason injuries. British journal of sports medicine. 2002;36(6):436–441.

27. Morgan N, Weston M, Nevill A. Seasonal variation in body composition of professional male soccer players. Journal of Sports Sciences. 2005.

28. Bangsbo J, Mizuno M. Morphological and metabolic alterations in soccer players with detraining and retraining and their relation to performance. Science and Football: London, UK. 1988.

29. Carling C, Orhant E. Variation in body composition in professional soccer players: interseasonal and intraseasonal changes and the effects of exposure time and player position. The Journal of Strength & Conditioning Research. 2010;24(5):1332–1339.

30. Dauty M, Collon S. Incidence of injuries in French professional soccer players. International journal of sports medicine. 2011;32(12):965–969.

31. Leventer L, Eek F, Hofstetter S, Lames M. Injury patterns among elite football players: a media-based analysis over 6 seasons with emphasis on playing position. International journal of sports medicine. 2016;37(11):898–908.

32. Della Villa F, Mandelbaum BR, Lemak LJ. The Effect of Playing Position on Injury Risk in Male Soccer Players: Systematic Review of the Literature and Risk Considerations for Each Playing Position. American journal of orthopedics (Belle Mead, NJ). 2018;47(10).

33. Hecksteden A, Kellner R, Donath L. Dealing with small samples in football research. Science and Medicine in Football. 2021;epub ahead of print; DOI: 10.1080/24733938.2021.1978106.

34. Batson S, Greenall G, Hudson P. Review of the Reporting of Survival Analyses within Randomised Controlled Trials and the Implications for Meta-Analysis. PLoS One. 2016;11(5):e0154870.

35. Della Villa F, Hagglund M, Della Villa S, Ekstrand J, Walden M. High rate of second ACL injury following ACL reconstruction in male professional footballers: an updated longitudinal analysis from 118 players in the UEFA Elite Club Injury Study. Br J Sports Med. 2021;55(23):1350–1356.

36. Arnason A, Sigurdsson SB, Gudmundsson A, Holme I, Engebretsen L, Bahr R. Risk factors for injuries in football. The American journal of sports medicine. 2004;32(1_suppl):5-16.

37. Hägglund M, Waldén M, Ekstrand J. Injury recurrence is lower at the highest professional football level than at national and amateur levels: Does sports medicine and sports physiotherapy deliver? British Journal of Sports Medicine. 2016;50:bjsports-2015.

