## Supplementary material for "The time course of injury-risk after return-to-play in professional football": "supplement" at line 168, and "supplementary document" at line 301

**Alternative data-processing 1: Randomly up-sampling on the individual level**

Here, the number of RTP episodes per player and season was balanced by random up-sampling on the individual level. Specifically, the RTP episodes originally recorded for a specific player within a season were amended by randomly drawing from this set until reaching the maximum number of RTPs per player in the corresponding season. For example, a maximum of 9 subsequent injuries from individuals was found for season 2014/15. For this season, injury records of players who sustained less than 9 subsequent injuries were amended by randomly drawing from the originally observed RTP episodes of the respective player within the 2014/15 season. The number of up-sampled injury records at individual level can be found in Figure S-3b.

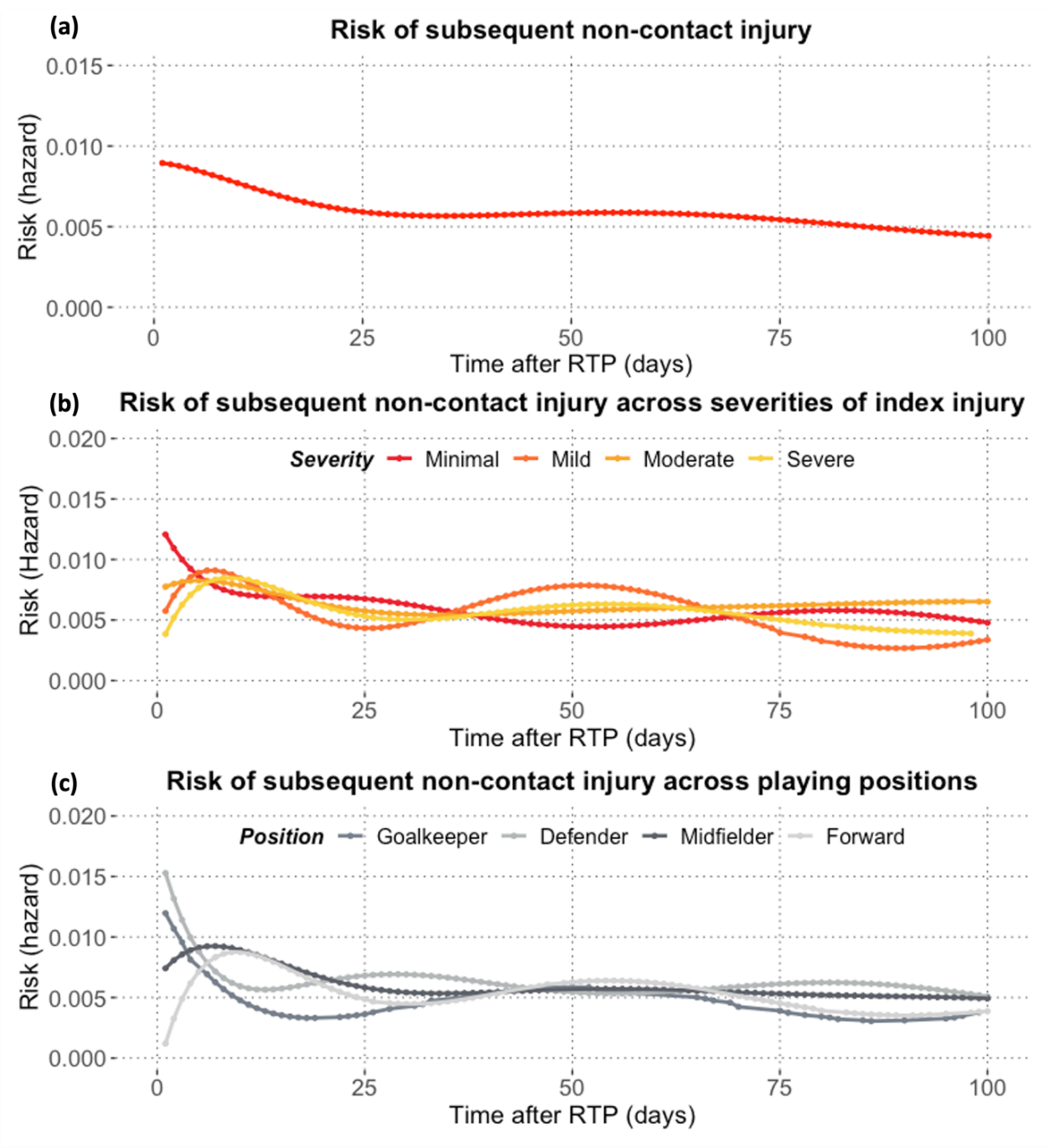

**Figure S-1.** Based on up-sampled dataset, the time course of a) non-contact injury risk after RTP; non-contact subsequent injury risk across b) severities of index injury, and c) playing positions.

**Alternative data-processing 2: Including only the first RTP of each player within each season.**

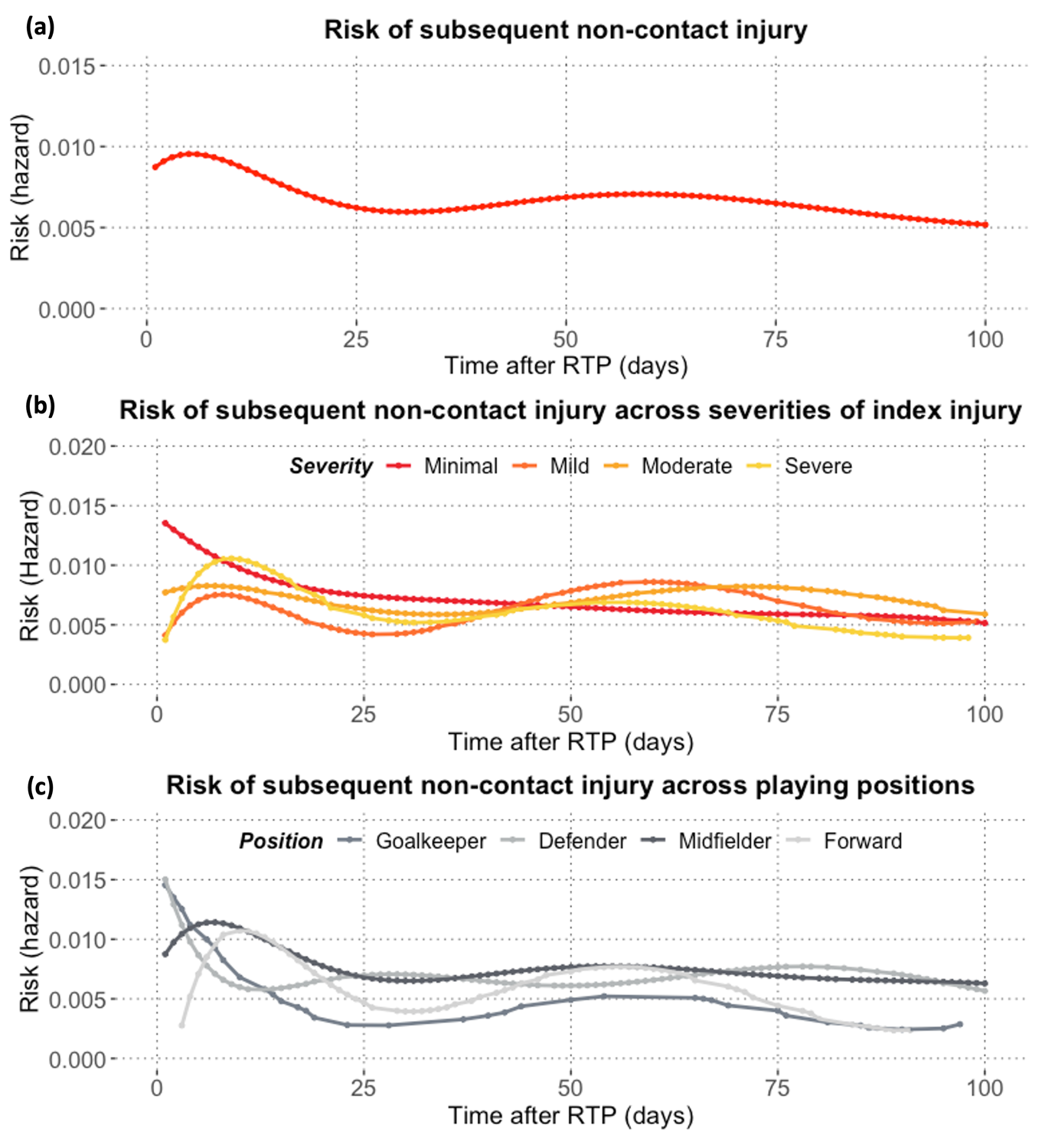

**Figure S-2.** Based on the first RTP of each player within each season, the time course of a) non-contact injury risk after RTP; non-contact subsequent injury risk across b) severities of index injury, and c) playing positions.

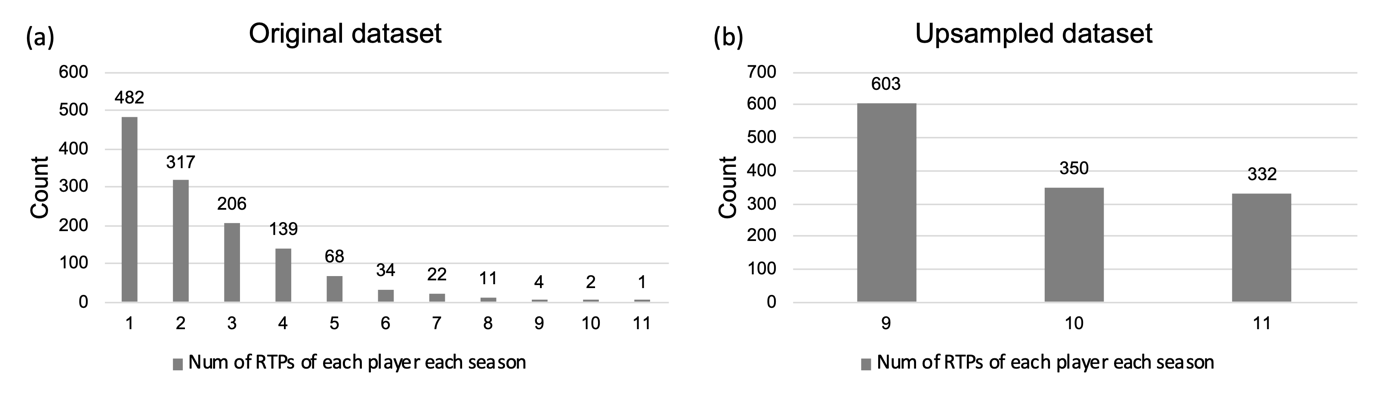

**Figure S-3.** The number of RTPs of each player within each season in the (a) original dataset; and (b) up-sampled dataset.

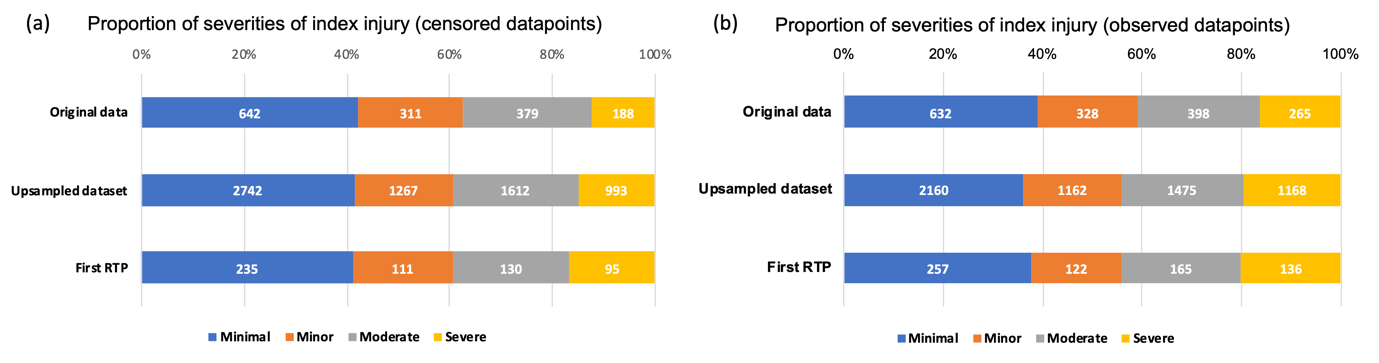

**Figure S-4.** Distribution of severities of index injury in (a) censored; and (b) observed data points, across datasets.
